# Long-term experience operating CDC recommended 5 air changes per hour in a K-5 elementary school using HEPA and MERV 16 Do-It-Yourself (DIY) portable air cleaners

**DOI:** 10.1101/2022.11.05.22281734

**Authors:** Devabhaktuni Srikrishna

## Abstract

On May 12, 2023 CDC recommended 5 air changes per hour (5 ACH) and in July 2021 California (CDPH) recommended 6 to 12 ACH to reduce long-range, aerosol transmission of COVID-19 and other pathogens in classrooms. EPA recommends MERV 13+ DIY air cleaners for temporary use during wildfires, and a recent EPA study reported inconsistent usage in homes due to excessive noise generated by the DIY air cleaners. Questions also remain about wear and tear including how long filters retain their filtration properties and need to be replaced. Herein we report real-world experience from daily usage of 47 HEPA and 60 MERV 16 DIY air cleaners in a California elementary school during two academic school years from spring 2021 through fall of 2023 across 16 classrooms, a library, an auditorium, a lunchroom, and in a hallway. Three to six purifiers were needed in classrooms to meet 6 to 12 ACH. Teachers reported noise generated by MERV 16 DIY purifiers on lowest fan speed as acceptable for classroom use. Filtration efficiency at 0.3 μm (most penetrating particle size) for DIY air cleaners with 5” MERV 16 filters used in the classrooms averaged 77% after six months (1st batch in February 2022) compared to 92% for newly installed filters (2nd batch in October 2022). Follow up testing on both batches of filters after an additional academic year (June 2023) showed only slight changes in average filtration efficiency. Portable air cleaners (HEPA and DIY) averaged and estimated 10 ACH (6-15 ACH) across the 16 classrooms demonstrating feasibility and unit economics of meeting CDPH targets per classroom for $200-$650 with DIY versus $600-$12,000 with the HEPA models used. In one 9000 cubic foot classroom with 7 air purifiers, air exchange rate was measured using ambient aerosols at 18 ACH from air purifiers (within 20% of ACH estimated based on CADR of purifiers) and 7 ACH from HVAC for a combined total of 25 ACH. Based on this long-term experience, specific recommendations for enhancing and improving CDC’s web page “Ventilation in Buildings” include: (1) recommended operation of MERV 13+ DIY at their low speed for low noise, cost-effective air cleaning (2) electro-mechanical safety especially in relation to power outlets (3) an open-source procedure known as the “spike test” using ambient aerosols to verify ACH in a room, like the Portacount for mask fit testing. Spike testing can become the basis for ACH certification or verification in any room without generating aerosol contaminants (e.g. salt water, smoke, tracers which may be unsafe or disallowed indoors).

## Introduction

A recent survey of N=106 California parents revealed their top concerns for classroom air quality are COVID-19 (50%) and respiratory viruses/bacteria in general (45%), although many also expressed anxieties about wildfire pollution and cognitive effects of bad air. Parents also complained it is not fair that certain school districts can afford to protect their students (air quality inequity), windows don’t open, CO2 levels are high, classrooms are overcrowded causing outbreaks especially Covid, there is a lack of transparency and attention to air quality from school administrators, lack of air purifiers, no HVAC in the school building, and lack of accommodations for the immunocompromised. 63% were unaware that California Dept. of Public Health (CDPH) recommends cleaning air 6 to 12 times per hour in classrooms, 79% said it was not clear from reading CDPH webpage that cleaning air 6-12 times per hour needed in classrooms, and 86% said clear and easy instructions is very important, 93% said low cost options and budgets are very important to include.

On May 12, 2023, for the first time CDC recommended a universal target of 5 air changes per hour (ACH) as the minimum air cleaning rate needed to reduce the risk of aerosol-transmitted infections like Covid-19 in occupied indoor spaces [13], such as offices and homes [14]. In recommending 5 ACH, CDC linked to a study conducted in 2021-2022 [18] in the Marche region of Italy. This region invested in ventilation systems in schools in advance of the Delta and Omicron waves during which transmission within classrooms was tracked with testing [17]. Analysis of classroom infection rates revealed they were over 80% lower in classrooms with 6 air changes per hour compared to schools without ventilation upgrades [16]. However, in 2022 in response to my query in JAMA [20], CDC experts [21] indicated more than 12 ACH may be necessary to stop spread of SARS-CoV-2 in some scenarios even though they were not aware of outbreaks in spaces ventilated at 5 to 6 ACH.

Currently few classrooms in California can meet the new CDC standard for indoor clean air of 5 air changes per hour (5 ACH) announced on May 12, 2023, less than the 2021 recommendation for schools by California public health for 6 to 12 ACH. An national survey by CDC shows few schools do air quality well, “None of the ventilation strategies was reported by > 51% of school districts” in CDC survey of 8,410 school districts (64.2%) representing an estimated 61.7% of enrolled public school students” [26]

Frequent air cleaning may not stop spread of SARS-Cov-2 at near-field (e.g. talking close by), but can potentially emulate far-field equivalent of N95 with 95% reduction of particles (20x) in a well-mixed room. Using 0.6 ACH as a baseline for unventilated rooms at least 12 ACH is required for far-field protection equivalent to N95 (95%), and 11-12 ACH was measured aboard three passenger airplanes in flight [19].

Portable air cleaners (also called “air purifiers”) and central HVAC systems are two valid ways to achieve the ACH targets necessary for infection control for Covid or many other harmful aerosols. Portable air cleaners that “recycle” the air in small to medium sized rooms may in many instances be cheaper (more cost-effective) than HVAC which consume high levels of energy and investment to transport, heat, and cool the “fresh” air from outside through existing ductwork that in turn may need to be upgraded to meet higher ACH targets demanded for infection control. While owners of some buildings can perhaps afford “fresh” air using the central HVAC to meet these higher ACH targets, “recycling” the air in each room with portable air cleaners, like plugging in lamps or fans in each room, can be more economical and scalable in terms of up front investment, energy consumption, and operating costs. In other instances, the outside air maybe polluted (e.g. during wildfires or from industrial pollution) making it undesirable unless well-filtered. A minimum 6 and preferably 12 air changes per hour is recommended by California Department of Public Health (CDPH) [5] to reduce and prevent airborne transmission of SARS-CoV-2 virus. In general, 6 ACH requires 100 cfm per 1000 cubic feet of space, and 12 ACH is double that.

Many HEPA purifiers sold in the United States are tested for clean air delivery rate (CADR) by the Association of Home Appliance Manufacturers (AHAM). Multiple research groups have similarly evaluated the effectiveness of Do-It-Yourself (DIY) air cleaners in the lab confirming their effectiveness to remove aerosolized particulate matter if used correctly [1] [2] including with filters capable of high filtration efficiency at the most penetrating particle size of 0.3 μm [3]. Subsequently, both CDC [22] and EPA [23] conducted laboratory evaluations of MERV 13+ DIY HEPA purifiers and concluded that (if well-constructed) they can cost-effectively remove bioaerosols (0.3 μm to 0.65 μm) and wildfire particulates (PM 2.5) respectively.

HEPA purifiers that deliver useful CADR on their highest fan speeds may be rendered much less effective for COVID-19 prevention when running on their lower fan speeds or auto speed, and end-users may forget to turn up their fan speeds to maximum.

However, EPA and CDC recommend DIY air cleaners only as a temporary alternative to commercially available air cleaners [13] [15]. An EPA study presented at the annual American Association of Aerosol Research (AAAR) in 2022 reported inconsistent usage and dissatisfaction of DIY air cleaners in homes during wildfire season in California. The most common complaint being excessive noise generated [4], and one reason maybe because end-users were advised to set them at the highest speed and reduce speed if necessary. Furthermore if DIY air purifiers were to be used daily, questions still remain about how long the filters will last or how often they need to be replaced.

MERV 13+ DIY purifiers based on the well-selected box fans (e.g. Lasko) and filters (e.g. Lennox, Nordic Pure, 3M) as detailed in [3] is that they can be provisioned and used on the lowest fan speed to provide useful levels of CADR in a classroom, home, or business without excessive noise generation that cause them to be turned off in most situations. A practical advantage of low-speed operation is it eliminates the possibility of running on lower speed settings than needed to meet the ACH target, frequently an issue observed with HEPA purifiers.

To mitigate concerns of airborne spread of SARS-CoV-2 virus during the COVID-19 pandemic and transient wildfire pollution, both HEPA and DIY air purifiers were assembled/installed by parents and used daily by teachers and staff at an elementary school in California during the academic school year from spring 2021 through fall of 2023 (ongoing) in 16 classrooms (30’x30’x10’ or 9000 cubic feet each), a library, an auditorium, a lunchroom, and in a hallway as shown in Figure 1. These included four models of HEPA purifiers (A, B, C, D) and DIY air cleaners (with Lennox and Nordic Pure filters). To address the questions about usage and filter longevity of both HEPA and DIY air purifiers, herein we report the real-world experience operating them at the school and assess the “wear and tear” of daily usage on the filters through measurements of filtration efficiency using the techniques described in [3].

**Figure 1:**
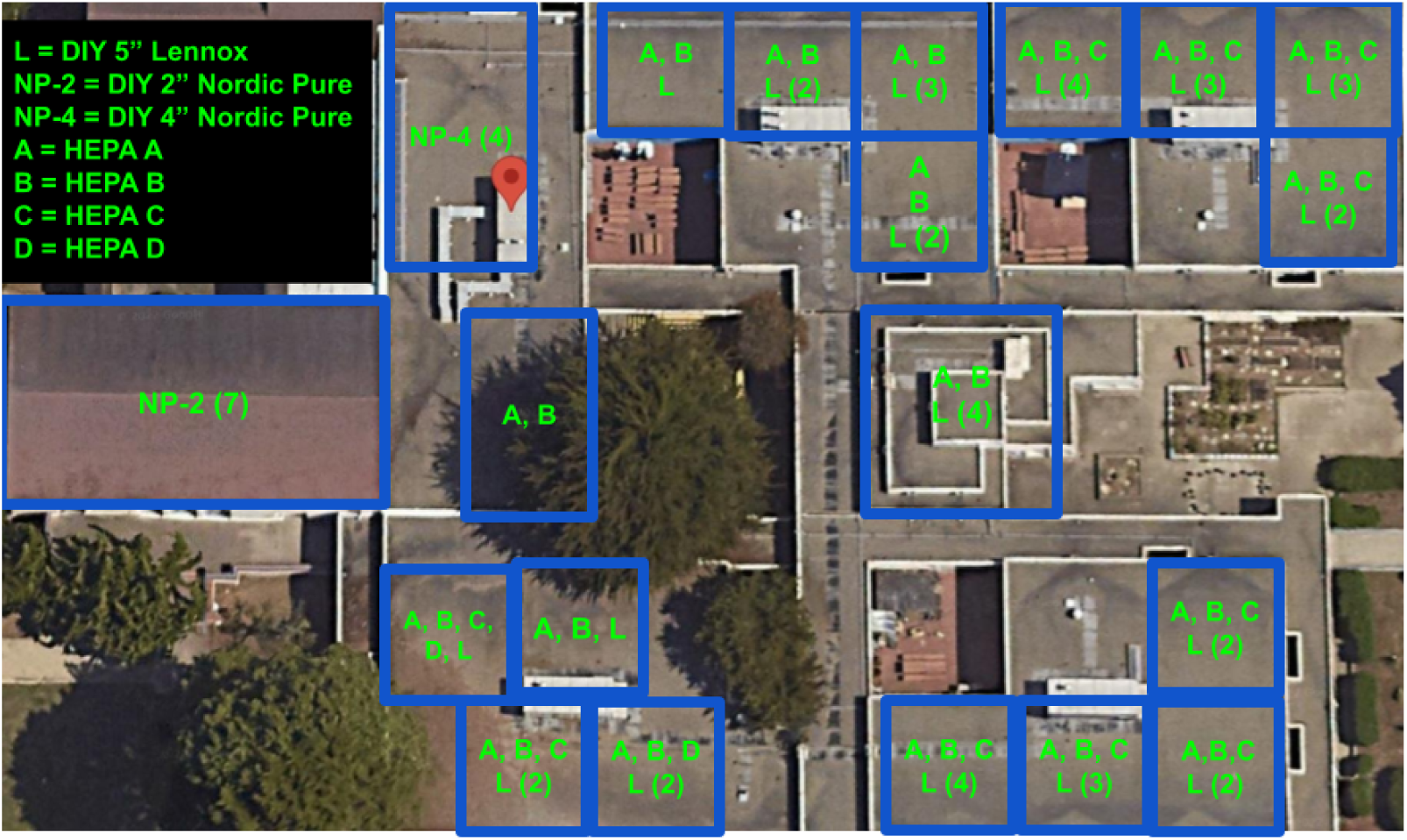
Map of school and placement of HEPA and DIY air purifiers.

Installation of HEPA A was conducted in Spring of 2021, HEPA B in Fall of 2021, and HEPA C and D in February, 2022. The DIY purifiers with two brands of filters operated at the low speed as illustrated in Figure 3. These DIY air cleaners were installed in or around February of 2022, with additional DIY purifiers (5” Lennox MERV 16) installed in or around October of 2022.

In Figure 2, the ACH from running air purifiers in each classroom is estimated using the CADR for each purifier from Table 1 and the volume of the classrooms (9000 cubic feet) ranging from 5.5 to 15.1 ACH, averaging 10.2 ACH across the 16 classrooms.

**Figure 2:**
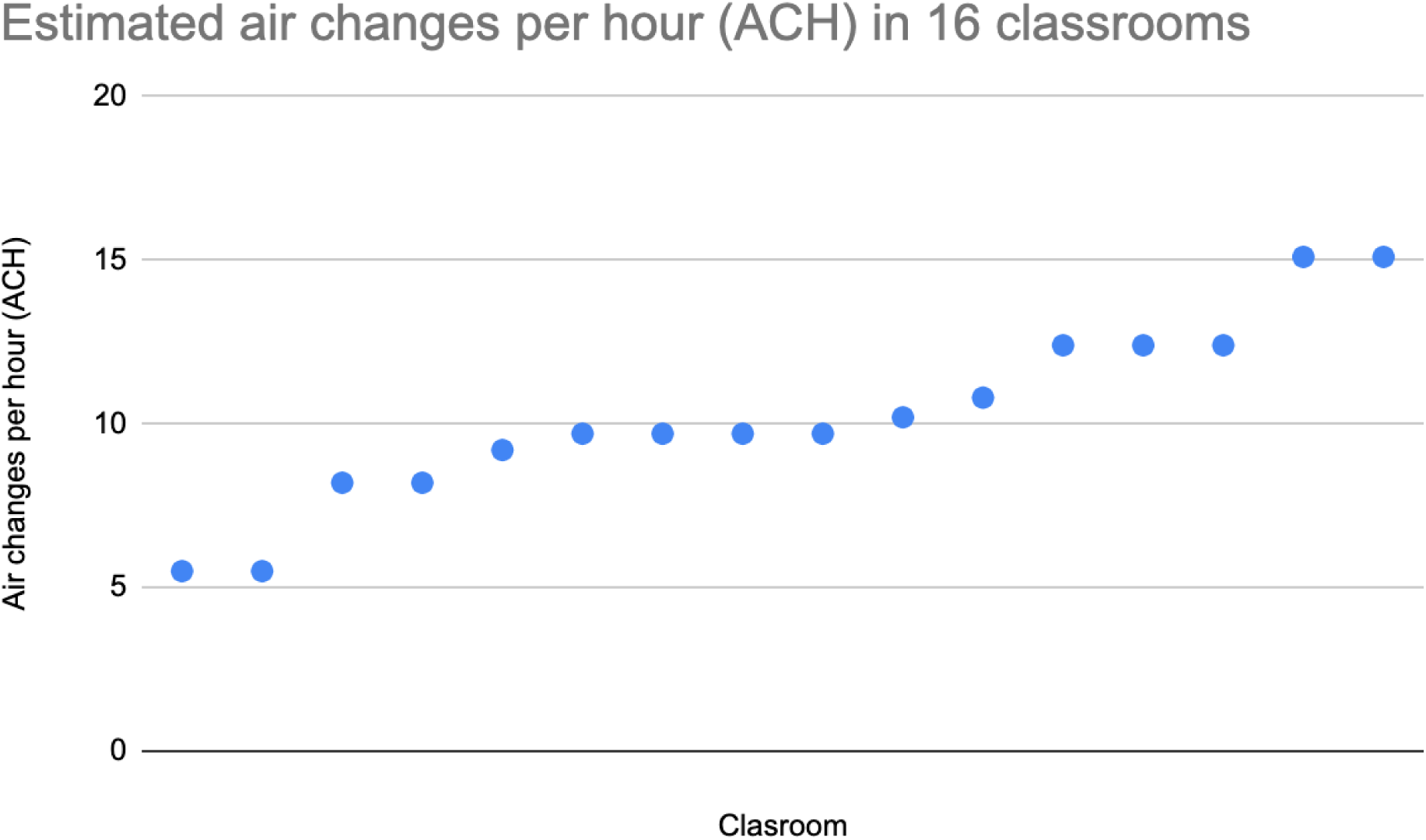
Estimated air changes per hour (ACH) in the 16 classrooms based on Table 1.

**Figure 3:**
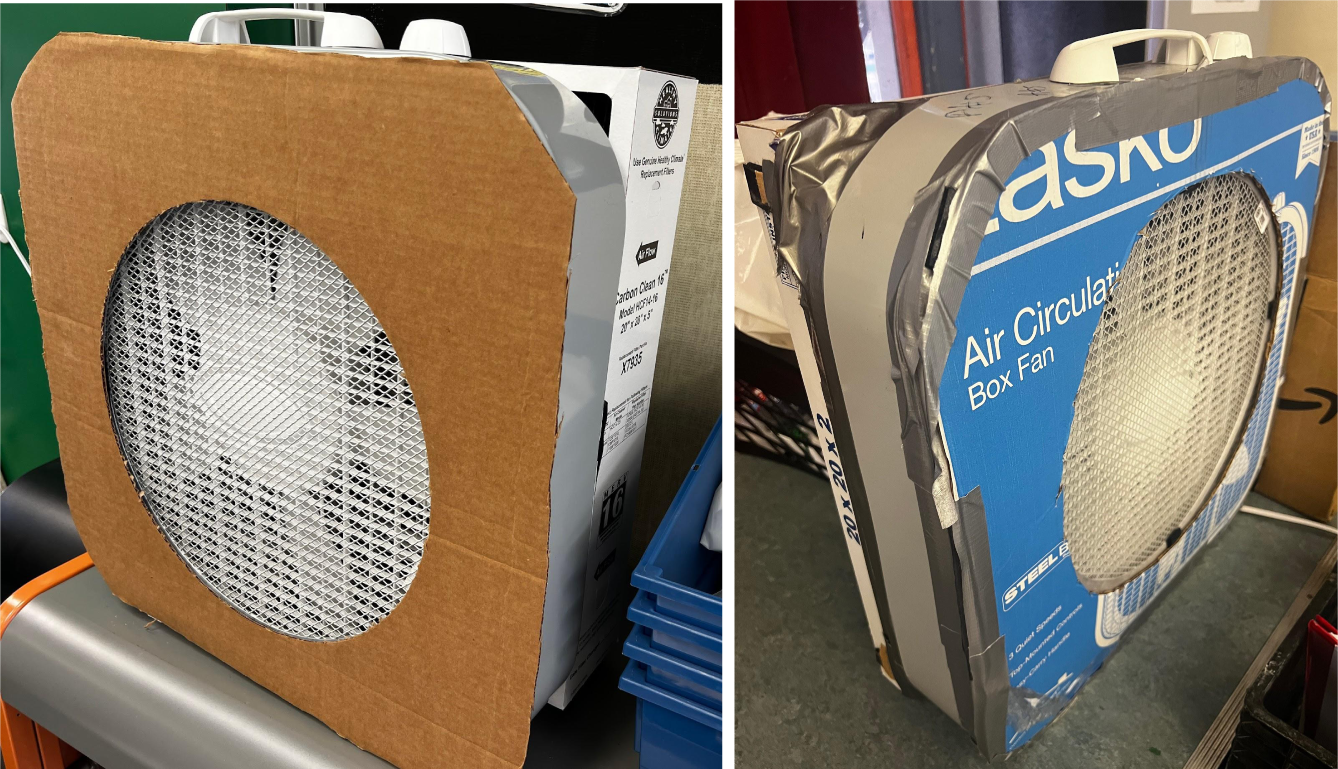
DIY air purifiers made from Lasko fans and 5” Lennox MERV 16 (left) and 2” Nordic Pure MERV 13 (right)

**Table 1:**
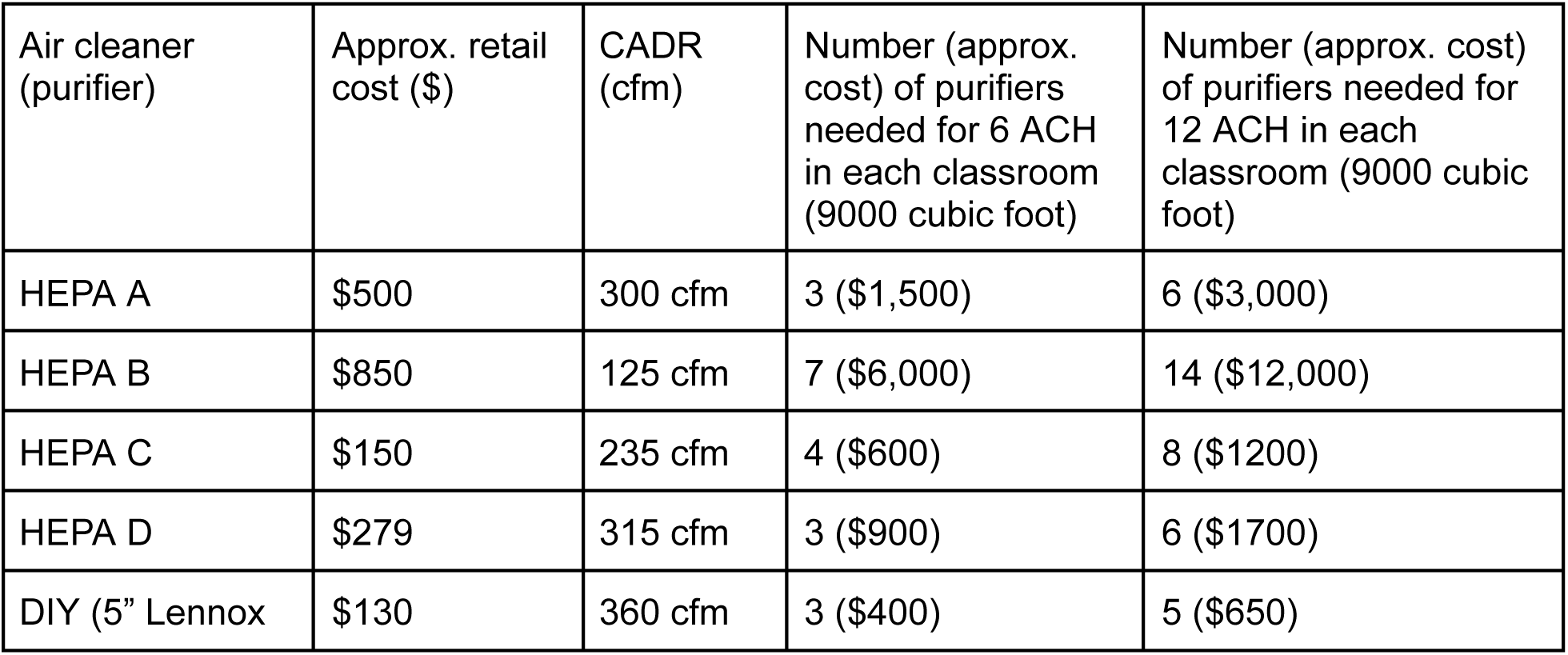
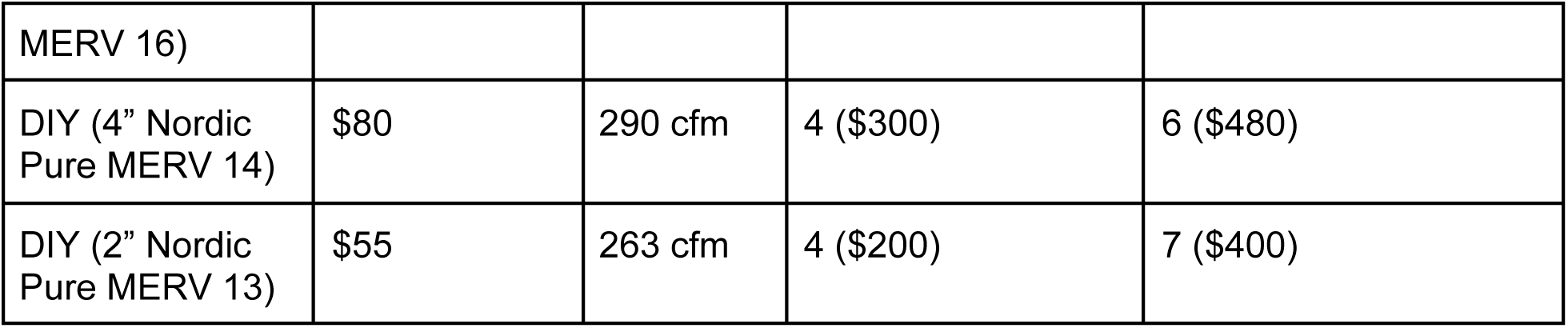
Filtration efficiency at 0.3 μm of HEPA and DIY air purifiers at the school.

Based on CADR testing results of DIY air purifiers at low speed [3] and to avoid the dissatisfaction of running them at high speed reported in [4], the teachers and staff were advised to use the DIY purifiers on the lowest speed only, and to run HEPA A on its maximum speed. The vast majority of teachers reported following those recommendations daily because the noise generated by those purifiers was tolerable in the classroom at these speeds. Whereas the vast majority of teachers also find HEPA B to be too noisy on its maximum speed and turn it down to the medium or low speed. The HEPA B are designated to be kept on 24×7, and all other purifiers are turned on/off by teachers and staff at the start and end of the school day respectively.

The DIY purifiers were made from Lasko box fans using a shroud and included an aluminum safety screen (pizza screen) in front of the fan since it is a K-5 elementary school with young children [6]. Each DIY purifier can be assembled in 8 minutes [7]. Each HEPA or DIY purifier in use at the school is estimated to deliver anywhere between 100 cfm to 400 cfm depending on the speed on which it is operated at. The progressive installation of air purifiers over time at the school reflects a desire to meet the minimum 6 and preferably 12 air changes per hour recommended by California Department of Public Health (CDPH) [5]. Additional DIY air purifiers are currently being installed in an effort to meet or exceed the preferred 12 ACH recommended by CDPH. For a 9000 cubic foot classroom in the school that is estimated to be over 6 ACH in most classrooms. At the lowest speed, the power consumption of Lasko fans was reported to be approximately 50 watts or 8 kilowatt-hours (kWh) per month if operating 8 hours per day in a school. At $0.10 to $0.25 per kWh operating the DIY purifier costs $1-$2 per month.

The number of HEPA or DIY units that could be placed was practically limited by available floor space and power outlets which varied from classroom to classroom. In some instances extra space could be found by rearranging items in the classroom, or power outlets could be expanded with power strips or expanders. In some classrooms there was a practical limit of three air purifiers.

## Methods

As described in [3], for each purifier measurement of filtration efficiency used a 7-channel optical particle counter (Temptop PMD 331) to count the ambient aerosol at input and output of the purifiers and measurement of airspeed at output using used an anemometer (Btmeter BT-100). In results below, all filtration efficiency measurements were based on recording 30 seconds of particle counts at input and output of the portable air cleaner or vent. Further research is needed, but published evidence suggests there is risk that significant quantities of SARS-CoV-2 virus as well as viable virus may be present in submicron aerosolized particles exhaled from people [9] [10] [11] [12]. In this study we compare the filtration efficiency of each filter at the 0.3 μm channel because it is known to be the most penetrating particle size [8], and in addition the 1 μm and 5 μm channels.

The method used to measure air changes per hour (ACH) in a classroom with a 7-channel optical particle counter (Temptop PMD 331) is described in [19]. The technique tracks the decay rate of ambient aerosols once the air purifiers are turned to arrive at an estimate of ACH and the “leak” of ambient aerosol particles into the classroom coming from the outside air ventilation system (doors and windows closed). The leak is in turn useful to estimate the rate of airflow from the ventilation system itself or through leaks from windows and doors in the room, separate from (in addition to) the air filtration provided by the purifiers within the room.

## Results [Fall 2022]

Table 2 below shows the average and standard deviation of the filtration efficiencies for each air purifier at 0.3 μm. Figures 4, 5, and 6 show the filtration efficiency for each of the purifiers in the school at 0.3 μm, 1 μm, and 5 μm respectively with x-axis representing each purifier ordered by the y-axis (efficiency). The measured efficiency for the Lennox filters is shown separately for those in use since February, 2022 (measured in or around October, 2022) and for new filters measured just after they were installed in October, 2022. Figure 7 shows the airspeed measured by an anemometer at the output of the air cleaner. The airspeed for each model of HEPA or configuration of DIY will vary based on its design or filter used, and airspeed is generally not directly comparable between different models or configurations.

**Figure 4:**
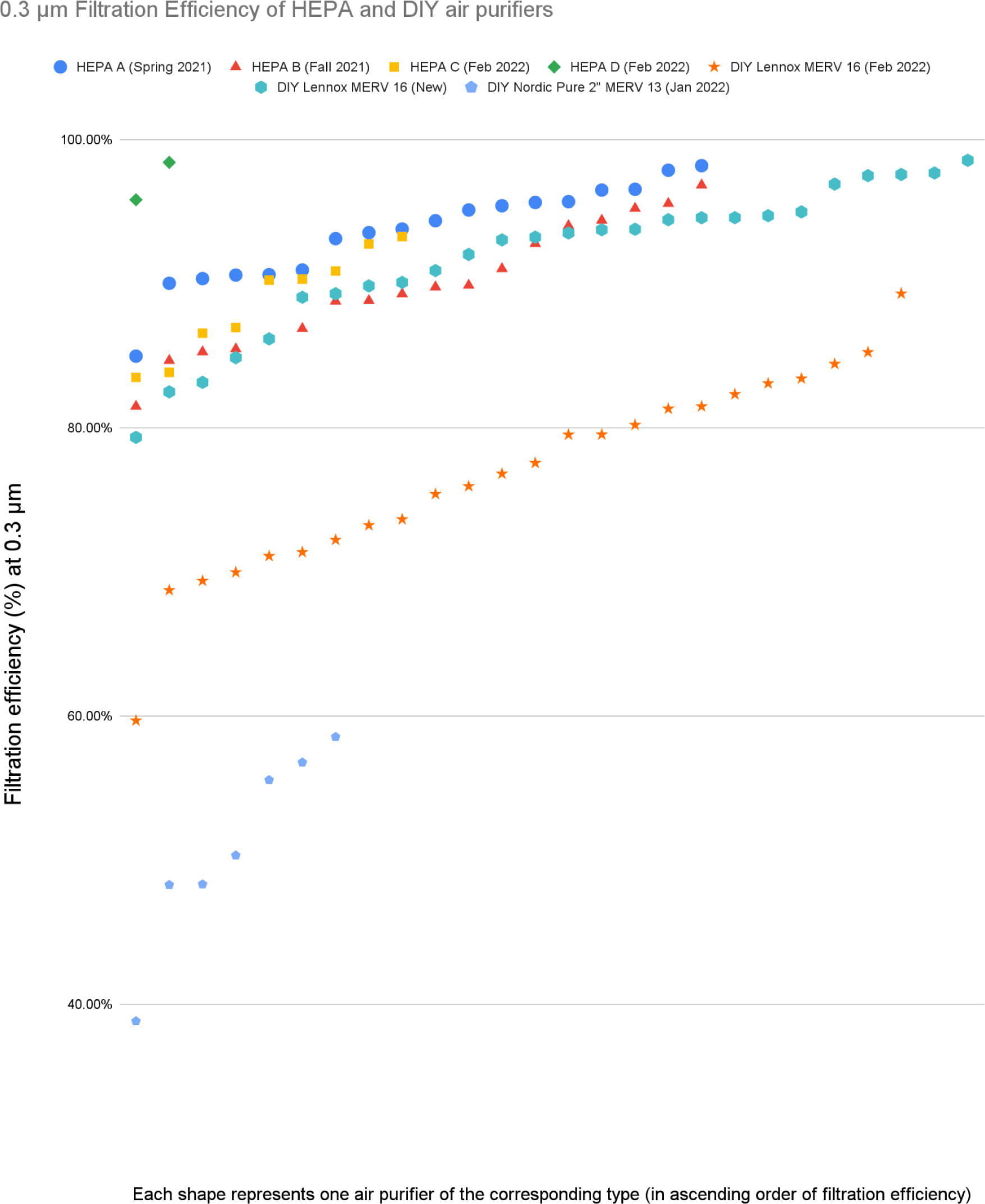
Filtration efficiency at 0.3 μm of HEPA and DIY air purifiers at the school.

**Figure 5:**
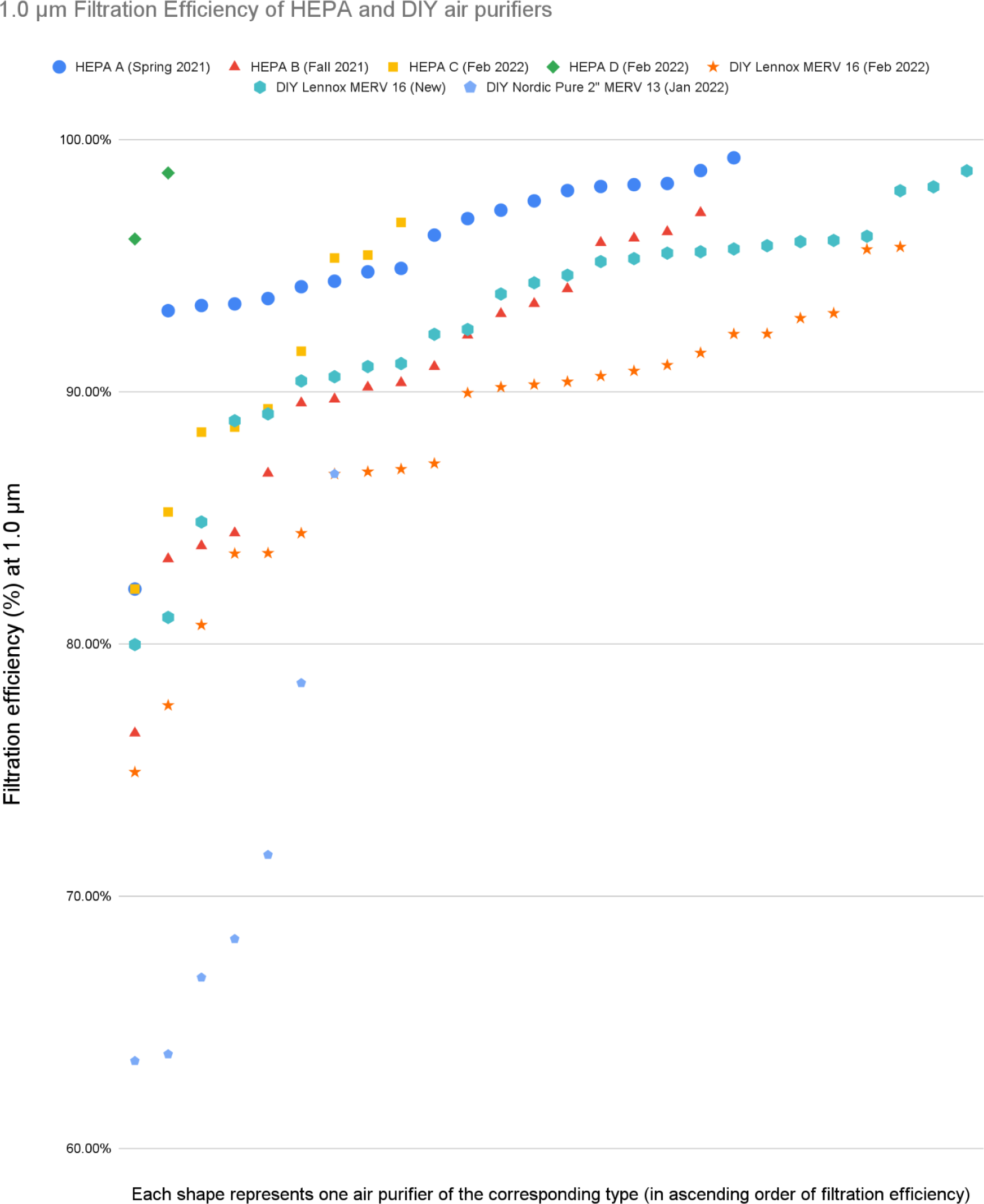
Filtration efficiency at 1.0 μm of HEPA and DIY air purifiers at the school.

**Figure 6:**
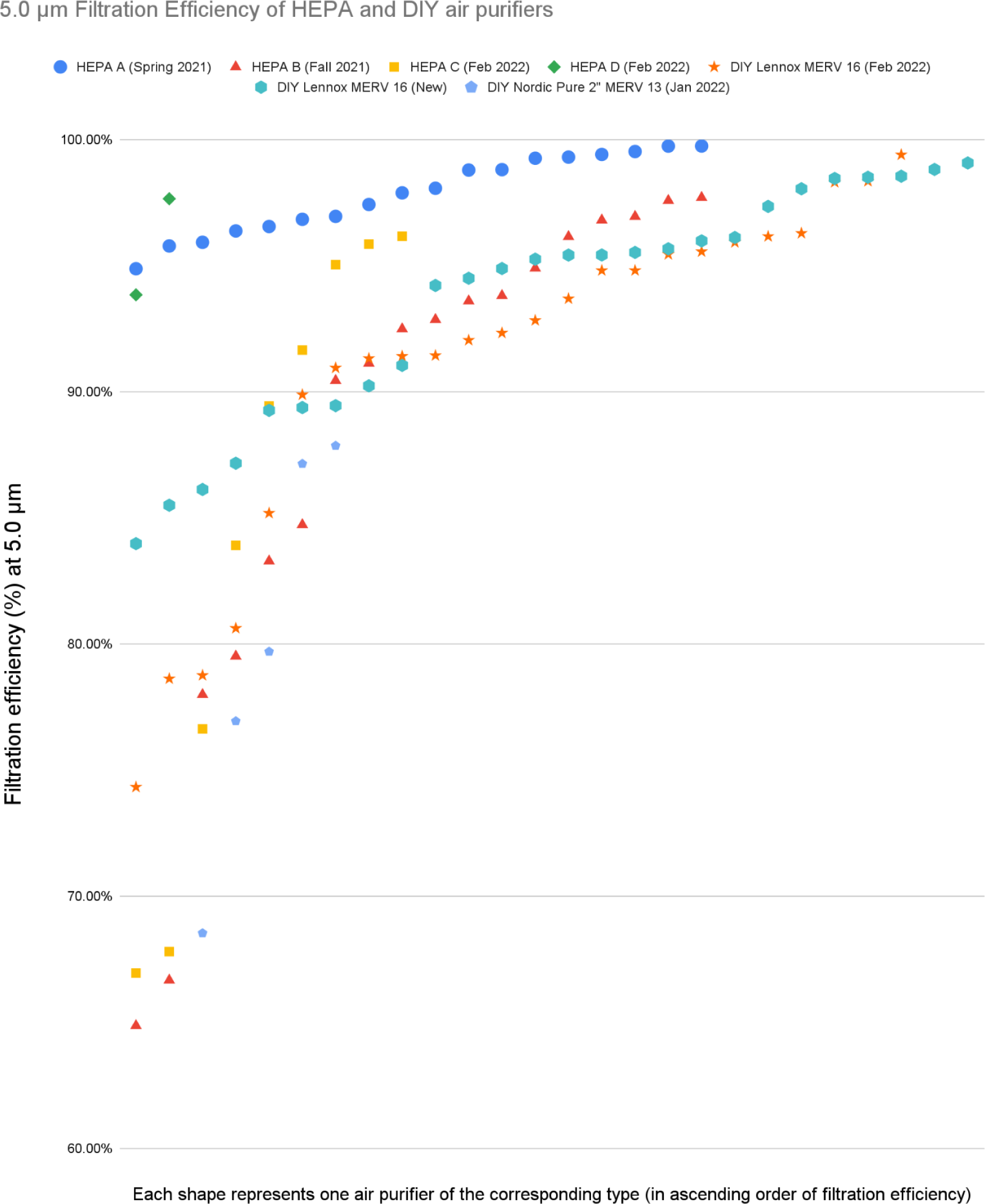
Filtration efficiency at 5.0 μm of HEPA and DIY air purifiers at the school.

**Figure 7:**
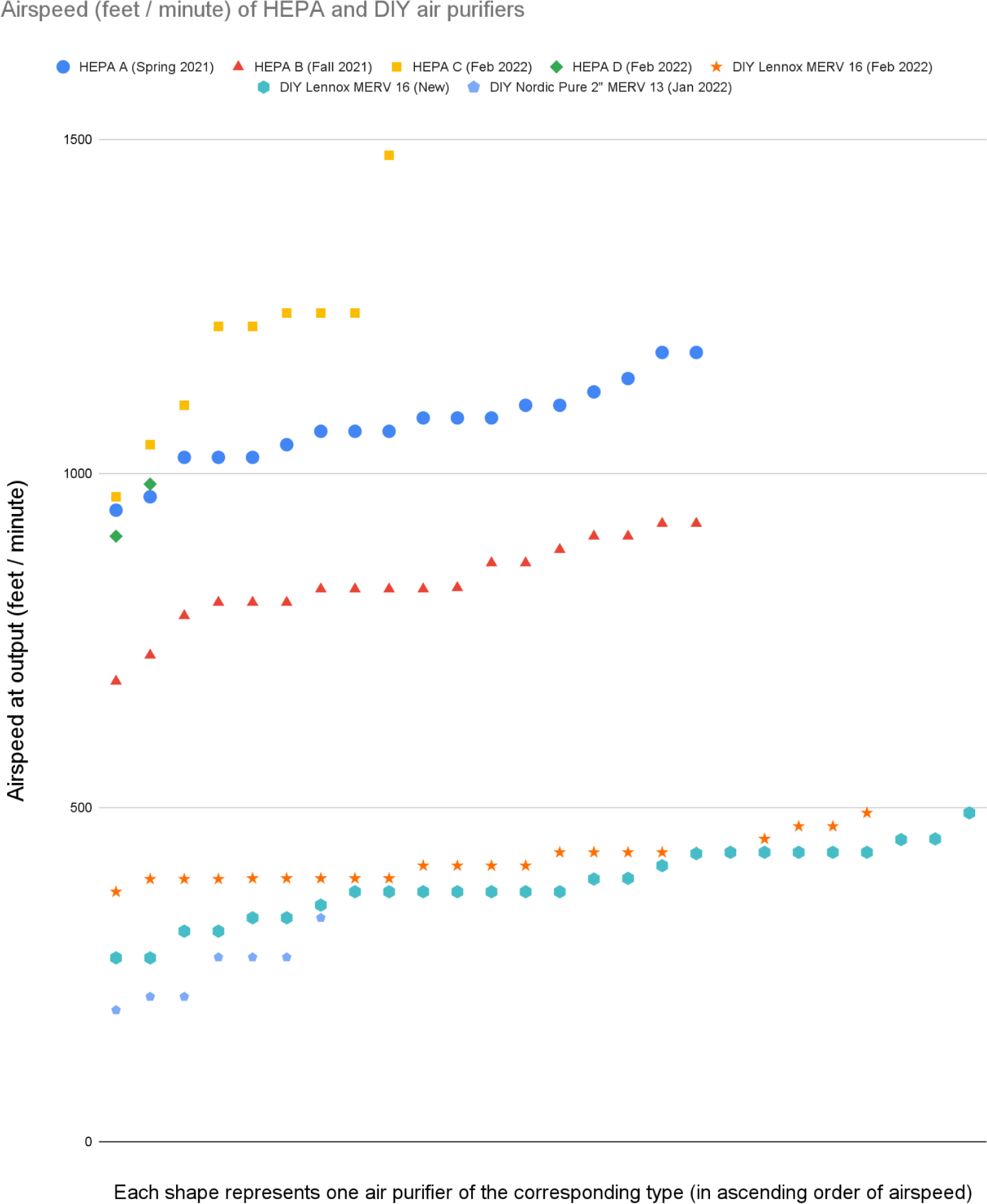
Airspeed of HEPA and DIY air purifiers at the school.

**Table 2:** Filtration efficiency at 0.3 μm of HEPA and DIY air purifiers at the school.

| Air Purifier | Count | Average | Standard Deviation | Minimum | Maximum |
| --- | --- | --- | --- | --- | --- |
| HEPA A | 18 | 93.51% | 3.38% | 84.96% | 98.18% |
| HEPA B | 18 | 89.79% | 4.34% | 81.48% | 96.83% |
| HEPA C | 9 | 88.69% | 3.62% | 83.49% | 93.26% |
| HEPA D | 2 | 97.11% | 1.83% | 95.81% | 98.40% |
| DIY Lennox MERV 16 (Feb 2022) | 24 | 76.87% | 6.71% | 59.68% | 89.30% |
| DIY Lennox MERV 16 (New) | 26 | 91.76% | 5.08% | 79.33% | 98.55% |
| DIY Nordic Pure MERV 13 | 7 | 50.95% | 6.76% | 38.83% | 58.55% |
| DIY Nordic Pure 4" MERV 14 (Jan 2022) | 4 | 62.38% | 4.42% | 57.90% | 66.35% |

**Table 3:**
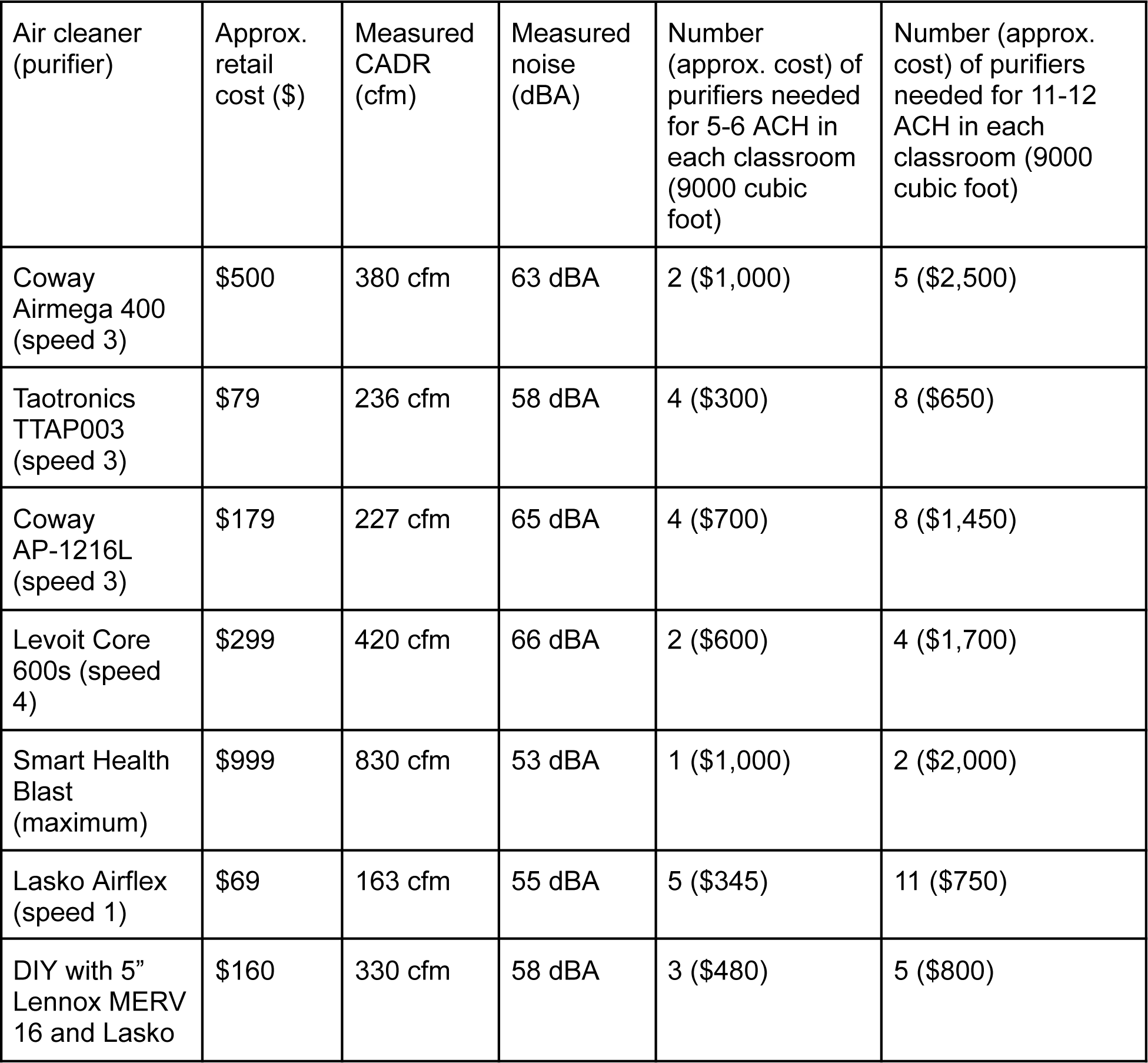
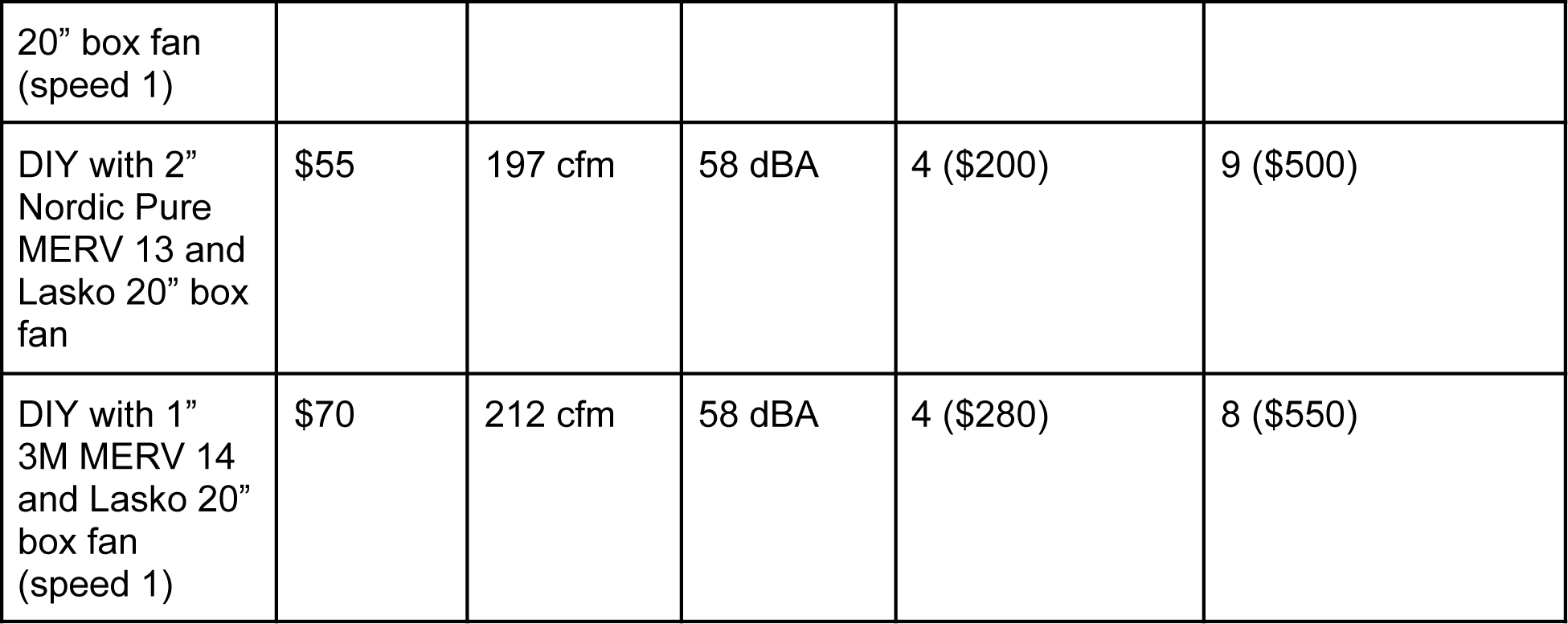
Approximate cost for 6 and 12 ACH using HEPA and DIY air purifiers.

## Results [Summer/Fall 2023]

In Figure 9, the filtration efficiencies (0.3 μm, most penetrating particle size) of the same filters measured in October 2022 are contrasted to longitudinal measurements of these filters in June, 2023 after a full academic year of daily usage. The average filtration efficiency of the batch of filters installed in February 2022 changed from 77% in (October 2022) to 74% (June 2023), and of filters installed in October 2022 changed from 93% in (October 2022) to 91% (June 2023). In contrast to Figures 4 through 7, each x-value in Figure 9 represents one filter instance comparing its filtration efficiency at different time points (October 2022 versus June 2023). Although some filters appear to have dropped, others appear to rise, possibly reflecting random variation in measurement of ambient aerosols. However, most filters experienced minimal change in filtration efficiency over the course of the past year.

**Figure 8:**
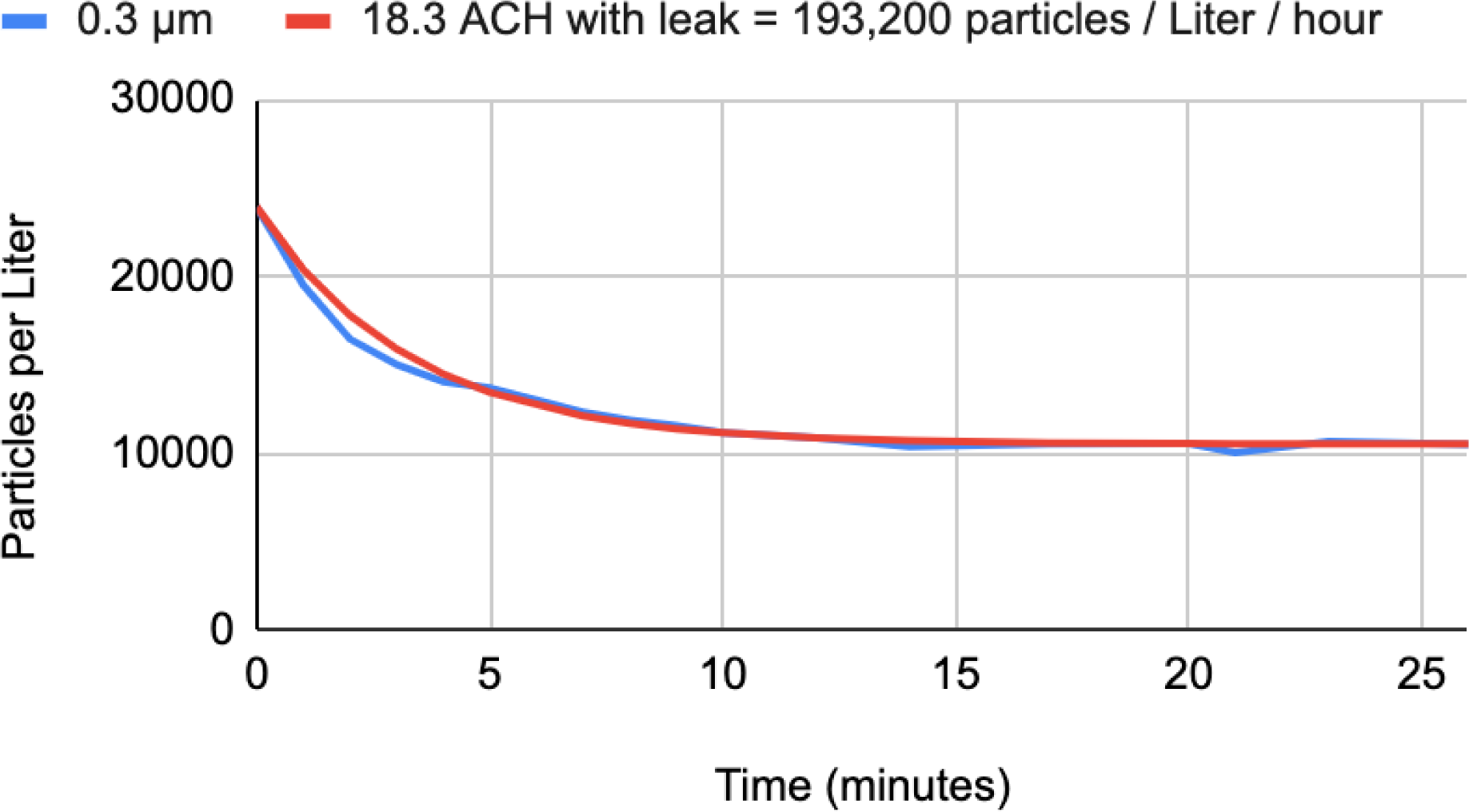
ACH measurement in 9000 cubic foot classroom with 7 air purifiers and HVAC.

**Figure 9:**
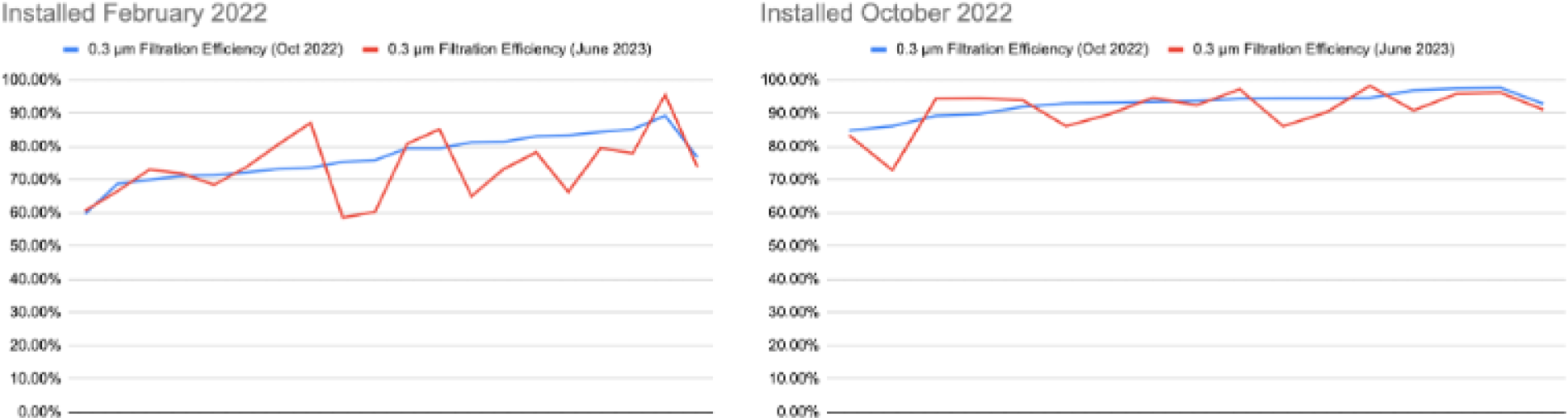
Filtration efficiencies filters measured in October 2022 versus June, 2023.

### ACH measurement using “spike test”

In one of the classrooms, the combined clean air delivery rate (CADR) of seven purifiers operating is estimated to be 2260 cfm from Table 1 (one each of HEPA A, B, C and four DIY purifiers with 5” Lennox MERV 16 filters). Based on this estimate, the ACH contribution from the seven purifiers would be 15 ACH (= 2260 cfm x 60 mins / hr / 9000 cubic feet).

Figure 8 shows the decay in particle counts measured at 0.3 μm in this classroom of size 9000 cubic feet (30’x30’x10’). The doors and windows were left open to “spike” the ambient aerosols prior to the test. Even though doors and windows were closed during the test, there were also four ceiling vents from the HVAC system that were supplying “fresh” air continuously into the room at an unknown airflow rate. In spite of “fresh” air from outside through the HVAC, the vents can be modeled as a “leak” of ambient aerosols into the room. Using the model in [19] based on particle decay rate, the ACH of the seven purifiers was estimated to be 18.3 ACH with a leak of 193,200 particles per liter per hour. The estimate of 15 ACH based on Table 1 is within 20% of the measured ACH of 18.3.

Just before the spike test, the 0.3 μm particle counts at the output of the HVAC vents were measured at 26,214 +/- 1,133 per liter (3 measurements), significantly lower than outside particle count measured at 59,466 per liter indicating the HVAC is likely filtering the outside “fresh” air. If the leak is mostly due from HVAC (not from closed doors or windows), the airflow rate in cubic feet per minute can be estimated by dividing the aggregate particle leak rate into the classroom (per 9000 cubic feet) by the concentration of particles per cubic foot measured at the output at HVAC vents (using appropriate conversions between liters to cubic feet and minutes to hours). This works out to be 1,100 cfm = (193,200 particles per liter per hr) X (28 liters per cubic ft) X (9000 cubic ft / 60 mins per hr) ÷ (26,214 particles per liter X 28 liters per cubic ft). In a 9000 cubic ft classroom 1,100 cfm delivers 7.3 ACH.

Similarly, a measurement of 6 ACH was verified in an 8000 cubic foot classroom (28’ x 24’ x 12’) with 3 MERV 15 DIY air purifiers at a nearby school which lacks any HVAC. The classroom was running two DIY air purifiers with 5” MERV 16 filters and one DIY air purifier with a 5” MERV 13 filter placed on the floor and constructed according to [6]. The aggregate clean airflow rate is estimated to be 800 cfm, or 266 cfm per air purifier. Although the classroom meets the desired 6 ACH, the CADR contribution per purifier was lower than expected (> 300 cfm) potentially revealing sub optimal placement of air purifiers near tables and walls.

### Electro-Mechanical Safety

While not often discussed in the context of portable air cleaners, electro-mechanical risks are inevitably introduced by regular use of any type of air cleaners (HEPA or DIY). Air safety efforts need a neighborhood watch for electro-mechanical aspects of safety, “if you see something say something” so qualified people can fix it right away:

- Electro-mechanical risk: Since air purifiers are devices that plug into AC outlets, ensure safe installation/operation by consulting your local expert for electrical, mechanical, earthquake safety especially with multiple air purifiers in a room.
- Wear and tear: Air purifiers may malfunction or be damaged due to wear and tear. Purifier wear and tear needs to be regularly inspected and taken care of right away when they arise.

There are several scenarios for end-users and the point-person to keep an eye out for electro-mechanical safety to look out for including but not limited to:

- broken plugs
- burnt or broken power sockets
- any kind of malfunctioning in a fan
- frayed cables (maybe a rodent chewed it)
- a fan stops working even when plugged in
- a fan rotating too slowly
- a fan making strange noises
- water damage

Below are some actual examples experienced at the elementary school that needed to be dealt with by end-users and point-person.

- Plugs can break off and pose a shock risk: A solution is to inspect plugs regularly and minimize plugs on the floor where people step if possible wherever possible. In an old school building with electrical sockets on the floor of library (not walls) a HEPA provided by parents was plugged into the floor and operated daily for 2 years without safety events. Two years later it seems someone finally stepped on the plug and broke it. As shown in the photos the metal prong of the plug broke off and was then lodged in the socket, effectively a live wire sticking out of the floor. Luckily the librarian (end-user) noticed it and showed it to the point-person, who immediately recognized it as a live wire, and taped it over before someone happened to touch the live wire (using whatever tape was in the room at that moment). That was a stopgap. The school administration put in a work order right away and the school district staff fixed it next morning without delay. The district also unplugged their own purifier from the floor.

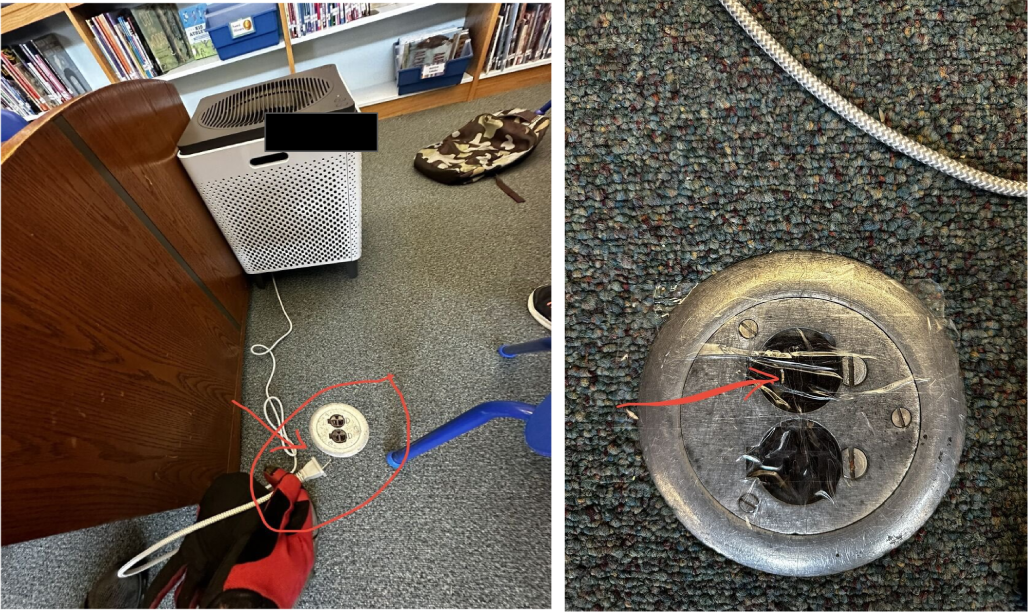
- Power socket extenders can overheat and/or catch fire: A solution is a fused extender with a circuit breaker. When too many things are plugged, if extenders exceed rated amperage they may melt or catch fire.

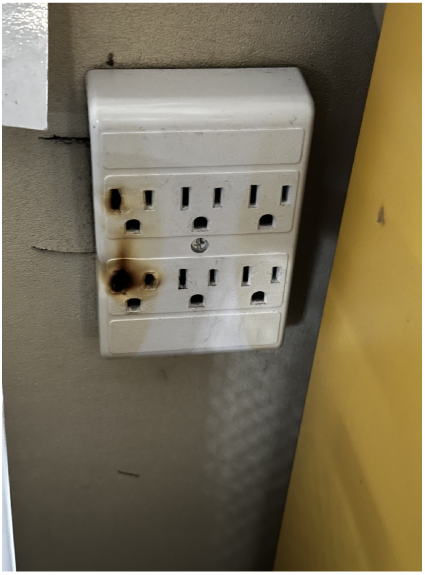
- Power-hungry devices may need to be plugged in directly: A big laptop charging box appears to have burnt out the above socket extender which draws a lot of power. It probably should have been plugged directly into the wall rather than through an extender. To provide outlets for the other devices, one solution is to plug the charging box into the wall directly, and then use a power strip with circuit breaker (instead of extender) in the second socket to supply the other devices.

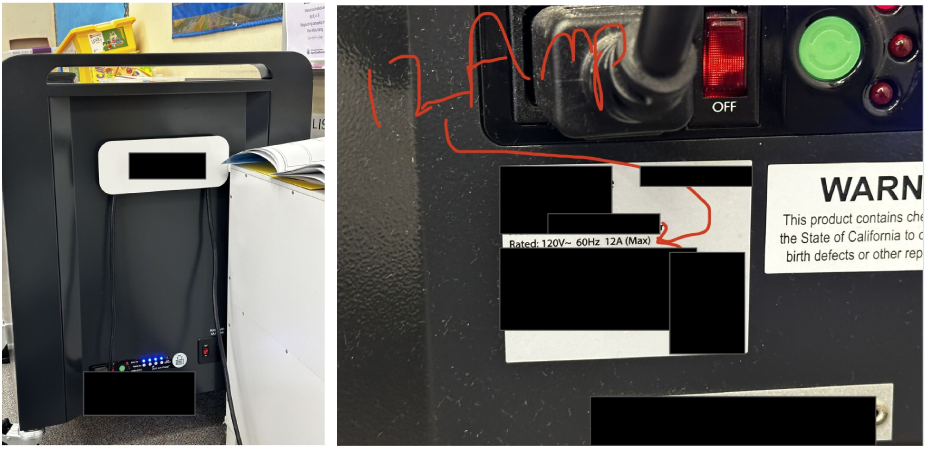
- Broken electrical sockets: An open socket was discovered while inspecting air purifiers before going back to school in Fall 2023. In such instances, it is necessary to call a qualified electrician. Once again the school administration put in a work order right and the school district fixed it next morning without delay.

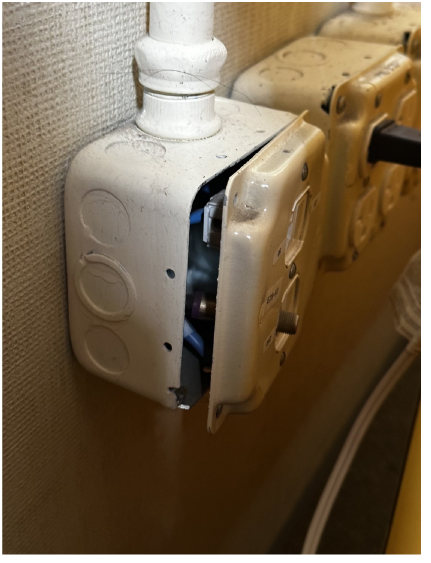

These electro-mechanical risks can be mitigated by taking a few simple but important precautions.

- Designate a point-person: A point person, someone who is frequently onsite, such as a volunteer parent, and capable of assessing electro-mechanical risks. The point-person should be designated for end-users such as teachers to raise concerns and immediately assess electro-mechanical safety. These include but are not limited to the scenarios described above.
- Tell end-users: “if you see something say something” which includes things like frayed power cords, malfunctioning fans, broken plugs, water damage, etc. and the scenarios described below but not limited to them.
- Do not DIY electrical failures: Once an electrical safety issue is spotted by the point person or end-user, do not attempt to fix electrical issues such as sockets, wires, etc. Refer electrical problems to qualified electricians. When it comes to electrical problems, it’s better to call in an electrician than Do-It-Yourself.

### HVAC Maintenance and Verification

In September, 2023 the elementary school experienced an intense wildfire smoke episode, during which the school instructed windows shut to prevent wildfire smoke from entering the classrooms. Using a handheld air quality meter (Temtop LKC-1000S+), the PM 2.5 particulate concentration (μg/m^3^) was measured inside at the HVAC vent and compared to outside in three classrooms. Each classroom was connected to a different air handling unit that serves a cluster of four classrooms each, upgraded in 2021 with filters that remove particulates.

As seen from Table 4, the air quality meter reveals varying levels of filtration from the HVAC in each classroom.

**Table 4:** Filtration efficiency in classrooms connected to different HVAC air handling units at the school during wildfire smoke event.

| Classroom | PM 2.5 at HVAC vent ($\mu\text{g}/\text{m}^3$) | PM 2.5 outside ($\mu\text{g}/\text{m}^3$) | Reduction via HVAC (%) |
| --- | --- | --- | --- |
| #1 | 38.5 | 39.9 | 4% |
| #2 | 19.3 | 30 | 36% |
| #3 | 17.4 | 47.2 | 63% |

Low-cost handheld meters can be useful to spot check the functioning of HVAC vents in the manner illustrated below. The vent in one classroom pumps air that is indistinguishable from the outside air, defeating the purpose of closing windows and running air purifiers to mitigate an intense wildfire smoke event. Based on this measurement, a work order was submitted by the school to replace the HVAC filters.

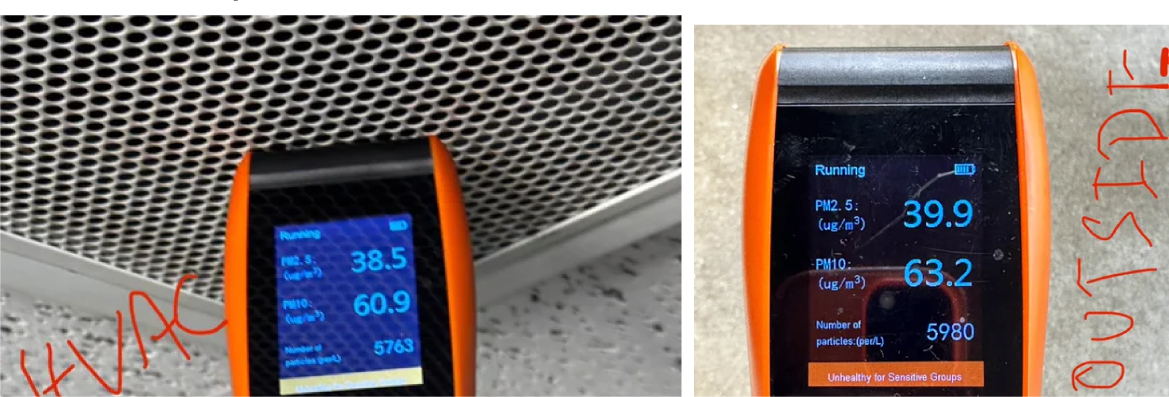

## Discussion

All purifiers are reported by teachers and staff to be in use daily during the school day, although purifier usage was not independently tracked through the school year on a daily basis (e.g. with a power consumption meter) it is possible some may not have been used consistently every day. However, the reduced filtration efficiency observed in the Figures 4, 5, and 6 between the new DIY Lennox MERV 16 measured at time of installation in October, 2022 (92% average) compared to measurements in October, 2022 of those installed in or around February, 2022 (77% average) is a reflection of these purifiers being used regularly if not daily since the latter were installed. Similarly the reduced filtration efficiency often below 95% and as low as 80% in some cases for many of the purifiers (HEPA A-C) in Figures 4-6 is also a reflection of “wear and tear” of frequent if not daily usage.

Three to six purifiers were needed in classrooms to meet California recommended 6 to 12 ACH. Deployment of HEPA and DIY at an elementary school in California [5]. The experience at a California K-5 elementary school demonstrates the feasibility of daily operation in 16 classrooms, library, auditorium, lunchroom, and hallway. Teachers reported noise generated by DIY purifiers (made of box fans) to be tolerable / acceptable for daily classroom use when run on low speed [25]. Filters in both DIY and HEPA purifiers held up for over six months of daily use, and in many cases over one year. Parents can be a local force for improving air quality in schools. Parents and PTAs tend to have close relationships with schools and are supplementing efforts by federal, state, and local governments to improve air classroom air quality.

Based on the measurement of reduced filtration efficiency after approximately six months of daily usage by teachers and staff, the filters in DIY air purifiers exhibited signs of “wear and tear” and are expected to last through one school year. Whereas the filters in HEPA purifiers may last longer. The HEPA models in use exhibited a wide variation in filtration efficiency at 0.3 μm among different units in the school ranging from almost 80% to well above 95%. This suggests some “wear and tear” on some of the filters which have been in use for one year or more, yet even at the lowest levels measured these efficiencies of HEPA purifiers remain useful to remove particulates in the room in a timely manner. If this trend continues and the filtration efficiencies continue to degrade at the same rate these filters perhaps may need to be replaced in a year or two.

The new Lennox MERV 16 used in DIY purifiers exhibited a much higher filtration efficiency at 0.3 μm (93% average) than DIY purifiers using the same model filters in operation since February, 2022 (77% average). Although the filtration efficiency was not recorded at the time of their installation in February, 2022 this difference may reflect “wear and tear” on the filters through the course of approximately half a school year. Subsequent measurements in summer of 2023 of these filter installed in Fall 2023 showed their filtration efficiencies held up by the end of one year (June, 2023) indicating most of the filters need not be replaced.

The 2”-4” Nordic Pure MERV 13-14 remain useful for filtering particles but the 2” MERV 13 exhibited reduced filtration efficiency at 0.3 μm (50% average) compared to measurements in [3] (> 60%), some of which were below 50% and may need to be replaced sooner.

Among 35 DIY air purifiers deployed in February, 2022 (Lennox and Nordic Pure) the fan on one of them stopped working, and another started making unusual noises. Both fans had to be replaced. Purifiers that might need their fan or filter replaced if their airspeed is much lower than others of the same model or configuration. Variation in airspeed can be seen among purifiers of the same model or configuration in Figure 7.

As verified in two classrooms of two schools (Figure 8 and Figure 10), tracking the indoor decay rate of ambient aerosol particles in the submicron range (0.3 μm) [19] can be used to validate the ACH of air purifiers (HEPA and DIY) separately from the ACH contribution from the HVAC system.

- In the 9000 cubic foot classroom estimated at 2260 cfm combined from 7 portable air cleaners, the exchange rate was measured at 18 ACH from air purifiers and 7 ACH from HVAC for a combined total of effectively 25 ACH. A high rate of particulate pollution measured outside at approximately 60,000 per liter was filtered down to approximately 25,000 per liter by the HVAC (“fresh” air) as measured at the vents and injected into the classroom at the rate of 7 ACH. The air purifiers further reduced this to approximately 10,000 per liter inside the classroom. During periods of extreme wildfire pollution, this measurement illustrates why the HVAC can counteract the filtering effect of the portable air cleaners if the HVAC supplying “fresh” air ends up injecting high rates of carcinogenic wildfire particles into the classroom. The air purifiers need to compensate for the pollution from outside air continued by the HVAC.
- In the 9000 cubic foot classroom estimated at 800 cfm combined from 3 DIY air purifiers, the air exchange rate was measured at 6 ACH. Although it exceeded the CDC target, the suboptimal placement of these air purifiers could potentially be improved to enhance the air change rate further.

**Figure 10:**
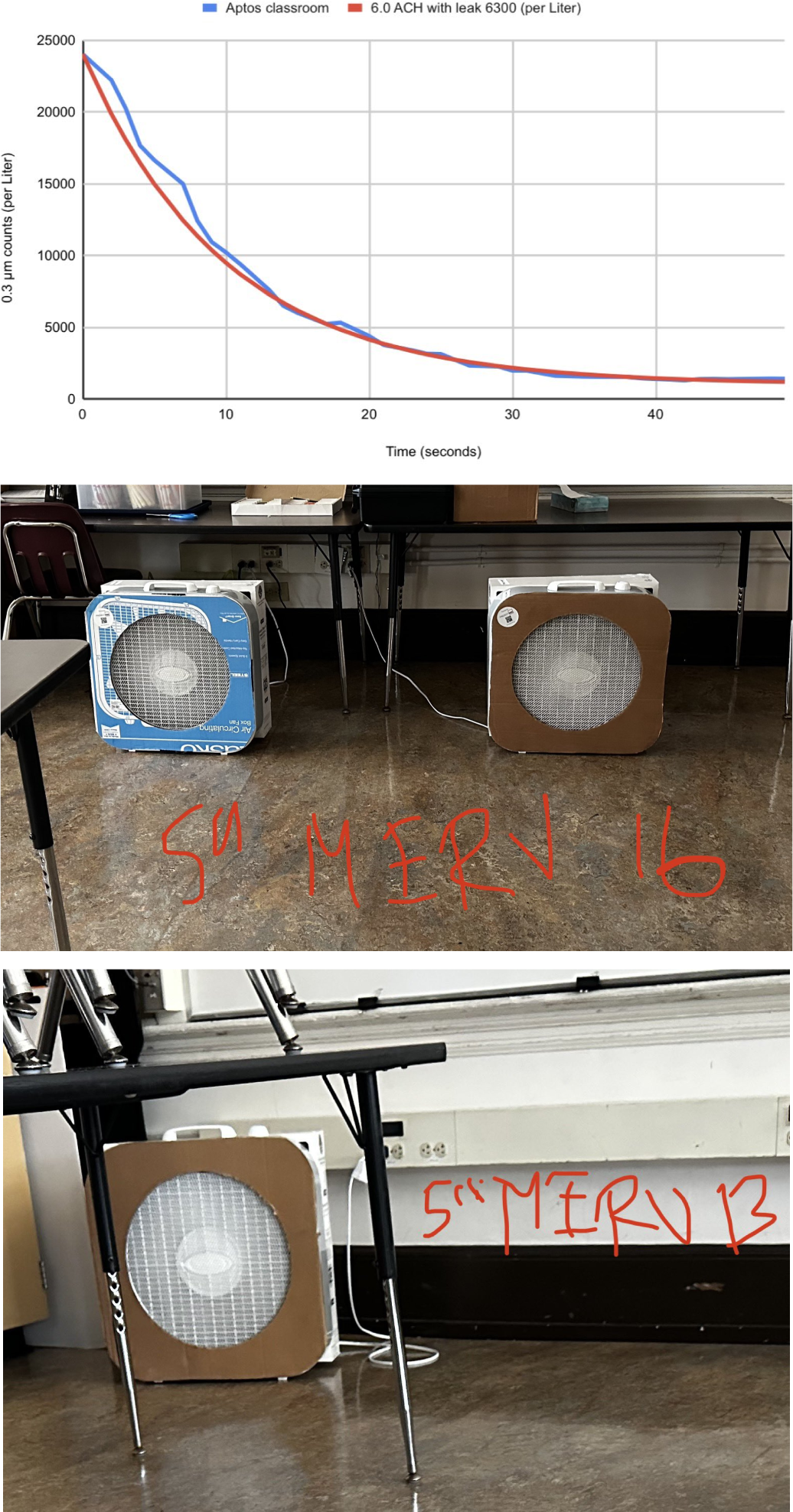
ACH measurement in 8000 cubic foot classroom with 3 air purifiers.

With or without central HVAC, portable air cleaners (HEPA and DIY) exceeded the level of air exchange (6-12 ACH) recommended by California Department of Public Health demonstrating feasibility and unit economics of meeting these targets using portable air cleaners per classroom for $200 to $650 with DIY versus $600-$12,000 with HEPA. Aside from the models in use at the school, a wide range of HEPA and DIY air purifiers with low-noise generation are available to reach the 5-6 and 11-12 ACH targets as tested in Table 2 of [19] and some representative models are shown in Table 3 below. The HEPA models are available through retail channels such as Amazon, Walmart, etc. DIY purifiers use the build procedure described in [6] [7] with parts available through retailers such as Amazon, Walmart, etc. It is important to note each room is different so any combination of air purifiers may result in different ACH than predicted from CADR, based on manufacturing variations, placement of purifiers, mixing conditions, speeds at which the purifiers are run, wear and tear, etc. The test procedure described using ambient aerosols (Figure 8) to verify ACH contributed by portable air cleaners and HVAC separately can be the basis for ACH certification or validation in classrooms and other types of rooms without generating aerosol contaminants (e.g. salt water, smoke, tracers) which may be unsafe or disallowed.

### Conclusion: Recommendations for enhancing CDC’s webpage “Ventilation in Buildings”

As discussed, parents can be a local force for improving air quality in schools. Parents and PTAs tend to have close relationships with schools and are supplementing the efforts by federal, state, and local governments. However to do so they need crystal-clear instructions from CDC on how to install, maintain, and verify the MERV 13+ DIY air purifiers to meet the CDC recommended target of 5 ACH in a safe and economical manner. Based on this long-term experience, specific recommendations for enhancing and improving CDC’s web page “Ventilation in Buildings” include: (1) recommended operation of MERV 13+ DIY at their low speed for low noise, cost-effective air cleaning (2) electro-mechanical safety especially in relation to power outlets (3) an open-source procedure known as the “spike test” using ambient aerosols to verify ACH in a room, like the Portacount for mask fit testing.

CDC recommendation of 5 air changes per hour provides a clear target for provision of ventilation in schools, in particular air filtration using portable air cleaners. However CDC’s webpage “Ventilation in Buildings” [13] provides a calculator for estimating the amount of clean airflow needed to meet the target, but does not offer a way to verify if the target is actually being met. As demonstrated above, low-cost the test methods are necessary to uncover if and when HVAC and portable air cleaners are failing to meet the target of 5 ACH, which can ideally be accomplished with an open-source test protocol provided on the same webpage [13] to verify and certify the ACH in any room. One candidate for the ACH test protocol is the “spike test” described relying on ambient aerosols, just like the Portacount for mask fit testing.

The CDC webpage [13] includes several statements regarding DIY air cleaners which could potentially be revised or updated based on our practical experience with DIY and HEPA at the elementary school, “While similar to commercially available HEPA air cleaners, the introduction of DIY air cleaners in a space can introduce new issues that need to be considered.” Each statement is included in Table 5 below with comments.

**Table 5:** Statements from CDC “Ventilation in Buildings” and recommendations for enhancement based on long-term operation at elementary school.

| CDC “Ventilation in Buildings” [13] | Recommendation based on school experience |
| --- | --- |
| “However, DIY air cleaners should not be used as a permanent, long-term solution for in-room air cleaning.” | MERV 13+ DIY air cleaners should be considered useful for more than temporary use. The case study demonstrates that MERV 16 DIY air cleaners can operate at scale for over one year, and potentially longer. |
| “Using multiple filters can reduce the resistance to air flow, allowing the fan to move more air when more filters are used in the design.” | Use of a better single filter can save space and achieve the comparable result as multiple filters. While multiple filters may improve the airflow compared to one filter of the exact same make/model, thicker and higher performing filters can improve the airflow compared to multiple thinner or lesser rated filters. |
| “Electrical cords for the fan need to be secured to prevent causing a tripping hazard.” | Tripping hazard is not unique to DIY, and can apply just as well to HEPA air cleaners. Furthermore a much greater risk arises from faults in use of sockets, expanders, and power strips and more generally regarding electro-mechanical risks as outlined in the section above on electro-mechanical safety. Few of these risks are necessarily unique to DIY and apply as well to HEPA. A section on electro-mechanical safety including topics discussed above would be useful to include. |
| “The DIY air cleaners can create enough noise to create difficulties communicating verbally. This often causes the units to be turned off or operated at a lower and less-effective fan setting, which defeats the purpose of having the air cleaner in the first place.” | This is not unique to DIY and applies to HEPA as well. The enabling factor of DIY is certain box fan models (such as Lasko) can provide ample airflow running at their lowest speed without excessive noise that is acceptable in classrooms [25]. Recommending use of such “low-noise” box fans models on their low speed (rather than high or medium) is likely to result in greater end-user satisfaction, and also avoids inadvertently turning them to a lower setting than necessary to meet the targeted ACH. |
| “Adequate amounts of duct tape should be used to prevent leaks during assembly.” | For single filter DIY builds, duct tape is unnecessary and Velcro suffices [3] |
| “However, the performance of well-assembled DIY units can be approximated by measuring the air exhausted from the fan (using an air flow hood or other similar equipment to measure air flow) and multiplying by the efficiency of the filters at capturing 1 to 3 micrometer ($\mu\text{m}$) particles. As an example, MERV 13 filters are at least 85% efficient at capturing particles in that size range.” | Filtration efficiency of filters of the same MERV rating varies greatly based on the make/model. For some MERV 13 filters, the filtration efficiency can dip significantly below 85% in a particle-size dependent manner, especially at the maximum penetrating particle size of 0.3 $\mu\text{m}$ (in particular see Figure 2-D of [24]). The filtration efficiency of the specific make/model of filter for DIY needs to be independently verified based on testing e.g. using an anemometer and particle counter [3]. |

## Data Availability

All data produced in the present study are available upon reasonable request to the authors

